# Knowledge, Attitude, and Practice of Birth Preparedness and Complication Readiness among women attending Antenatal Clinic in Enugu Metropolis

**DOI:** 10.1101/2024.10.06.24314964

**Authors:** Chisom Tony-Igwe, Udeani Kingsley, Udeichi Nnenna, Ugwu Elina

## Abstract

**Introduction:** High maternal mortality is a major problem in the developing world.

Ill birth preparedness and complication readiness is the major single most important cause of maternal mortality in developing countries in Africa, of which Nigeria is one of the populous. This study aims to assess the level of Birth Preparedness and Complication Readiness[BPCR] among pregnant mothers attending antenatal clinic in ESUTH Parklane - the teaching hospital in Enugu Metropolis.

**Methods:** This was a questionnaire based cross-sectional descriptive study on the knowledge, attitude and practice of birth preparedness and complication readiness among pregnant women who attend antenatal care clinic in ESUT Teaching Hospital Parklane.

**Inclusion criteria:**

1. Pregnant mothers
2. Those willing to participate
3. Those available at the time of data collection.

**Exclusion criteria:**

1. Those not present on the day of examination.
2. Those unwilling to participate.

**Results:** Our study showed that 81.8% of respondent has heard about birth preparedness and complication readiness.

The study showed that the highest number of respondent first heard about birth preparedness and complication readiness from antenatal services (61.1%), followed by health workers (20.8%) then social media (11.6%).

The respondent had a knowledge of at least one danger signs of which vaginal bleeding was ranking the highest in pregnancy (61.4%), childbirth (43.6%) and post partum (46.6%) followed by foul smelling vaginal discharge (10.6%), (6.8%), and (7.6%) respectively and other symptoms. A vast majority of the population felt that birth preparedness and complication readiness was important (93.2%), with many pointing out that they would preferred to come to antenatal care(89%) as against staying at home or going to work, the compliance level of these women to regular antenatal care visits is (94.7%) and many more complaint with routine drugs(92.8%).

91.3% are prepared for the birth of the child, with majority having happy emotions (83%) towards the pregnancy. Spouse support was (95.5%).

**Conclusion:** Knowledge of birth preparedness is very high among the study population.

High level of spouse involvement in our study population and compliance to antenatal visits and routine drugs, good antenatal services, stable financial income, family size and outcome of previous pregnancies affects level of preparation and complication readiness and should lead to good pregnancy outcome, improved family health, increase living status and reduced maternal and infant mortality rate.

## Chapter One

## Introduction

### Background Information

Being prepared is a very vital tool in all of life’s dealings. It is well known saying that “he who fails to plan, plans to fail”. This implies that a lack of adequate preparation for any given event or task deemed important may inadvertently lead to undesired results. Planning indeed is a basic prerequisite that ensures that life and all its activities go as is required in the area of health, not consciously planning for good health by adopting healthy lifestyles, proper feeding, and hygiene, good personal and environmental hygiene, making a healthy budget, going for regular checkups e.t.c may lead to unnecessary and unwarranted health catastrophes in the long run and ultimately untimely death. Individuals with chronic health conditions like diabetes, hypertension, epilepsy, and sickle cell disease are usually strictly counseled on careful living and adequate preparation for crises and emergencies.

Pregnancy periods are well-known as high-risk periods in a woman’s life as a double burden is placed on the vital organs like the heart, kidneys, liver, lungs, and system such as circulatory system leading to physiological anemia. The immune system is also affected leading to an immunocompromised state with subsequent susceptibility to infections. Psychological and emotional changes such as depression, irritability, excessive cravings for certain foods e.t.c also occur. Pregnancy is termed ‘term’ when it has been carried out m for 37 weeks-42 weeks. This is the period where labor and delivery occur and should be anticipated. Planning for delivery is highly imperative because pregnancy is a high-risk condition, especially in women with additional conditions like placenta previa, breech presentations, uterine abnormalities, multiple gestations, diabetes, hypertension, sickle cell, etc., and in those that have had previous miscarriages and previous c-sections.

The process of labor itself can lead to complications to the fetus like birth asphyxia from prolonged labor, head injury, and subsequent brain damage due to instrumental delivery, shoulder dystocia in macrosomic babies, meconium aspiration, and maternal complications such as retaining of the placenta and even maternal and child mortality. All these can be avoided or managed properly as the case may be if adequate birth preparedness is in place. Birth preparedness also ensures pregnant women are in the right psychological and emotional state of mind for handling delivery and labor pains among others.

Birth preparedness is the process of planning and preparation for delivery which ensures timely reach to professional delivery care when labor begins, anticipating the actions needed in case of an emergency and reducing obstetric delays when complications arise. A lot of emphasis is placed on preventing delays i.e delays in seeking care, delays in reaching care, and delays in receiving care by ensuring knowledge of obstetric danger signs, identification of skilled birth attendants, and the closest and most appropriate health care facility, knowledge of the expected date of delivery, signs of labor pain, plan for transportation to the facility for delivery or when complications arise saving up money to pay for care and other resources identification of potential blood donor or decision maker in case of emergencies, arrangement for a temporary caretaker for the family while the mother is away, counseling on exclusive breastfeeding. This way the pregnant woman is adequately prepared for expert management on the day of labor and prompt intervention if any complications surface.

High maternal mortality is a major problem in the developing world. Globally, about 287,000 mothers die each year because of problems related to pregnancy and childbirth. About 99% of this occurs in developing countries; sub-Saharan Africa [56%] and South Asia [29%] account for 85% of the global burden of maternal death. Nigeria is unarguably the most populous country in sub-Saharan Africa and indeed in Africa as a whole and maternity rate is more than 15x higher slow progress in low and middle-income countries is explained by the low coverage of maternal health care and the 3 delays to health-seeking behaviors of mothers.

Ill birth preparedness and complication readiness therefore are the single most important cause of maternal mortality in developing countries of which Nigeria is one of the populous. The issue of birth preparedness and complication readiness thus cannot be overemphasized in ensuring maternal and child health is preserved during prenatal, perinatal, and postnatal periods.

### Problem Statement

Safe motherhood is an initiative of the United Nations (UN), launched in 1987 to ensure that women go through pregnancy and childbirth safely and give birth to healthy children, reinforced by maternal mortality reduction in the MDGs of 2000–2015. Yet every year, hundreds of thousands of women die or suffer serious complications from pregnancy and childbirth. Safe motherhood begins before conception with proper nutrition and a healthy lifestyle. Planned pregnancy, appropriate prenatal care, prevention of complications when possible, and early and effective treatment of complications are all essential to maternal care.

The idea of birth preparedness and complication readiness is a safe motherhood initiative to plan for a normal birth and anticipate the action needed in case of emergencies. Preparing for birth ranges from educating the pregnant woman, saving [money], getting the necessary items required for delivery, finding the nearest center, means of transportation, and awareness to neighbors.

Maternal death is a human tragedy and a violation of women’s right to life. Globally, the maternal mortality ratio has been reported to be 216 per 100,000 deliveries with 303,000 maternal deaths occurring in 2015. The majority (99%) of these deaths occur in developing countries with sub-Saharan Africa accounting for more than 66% (201,000 maternal deaths) of the global estimate of developing countries. Nigeria is one of the eighteen countries in sub-Saharan Africa that has a very high maternal mortality ratio (MMR) in 2015. It was estimated to be 814 (596-1180)/100,000 live births and it is higher in areas where there is poor access to skilled birth attendants.

The importance of birth preparedness and complication readiness cannot be overemphasized, as it can curb the major causes of maternal mortality such as severe bleeding (mostly bleeding after childbirth), infections (usually after childbirth), high blood pressure during pregnancy (pre-eclampsia and eclampsia), complications from delivery, and unsafe abortion.

Also, according to a Survey done in the United States of America, it was estimated that the total cost of nine maternal morbidity conditions for all pregnancies and births in 2019 was $32.3 billion from conception to five years postpartum, amounting to $8,624 in societal costs per birthing person.

Countries in the Subsaharan Region of Africa of which Nigeria is inclusive, are third-world countries with low standards of living and ever-increasing exchange rates in contrast to the stronger dollar, pounds, etc, the financial burden of maternal morbidity/mortality in this region is therefore monstrous.

Mental disorders such as postpartum depression are anxiety are also prevalent, as well as postpartum psychosis, postpartum Obsessive Compulsive Disorder, and Pregnancy-associated social phobia. These lead to problems during the pregnancy period and contribute to maternal morbidity/mortality.

However many women and the society in our immediate environment pay little attention to this aspect of childbirth thus leading to increased incidence and prevalence of high maternal, perinatal, and neonatal deaths.

According to a Cross-sectional, Multicentre study carried out in Benin Central Hospital and the University of Benin Teaching Hospital (UBTH) Edo State among 300 women on “the concept of birth preparedness in the Niger Delta of Nigeria”, only 38% of the respondents were aware of birth preparedness. Only 28.1% embraced birthpreparedness because of ease of child spacing, and only 23.7% embraced birthpreparedness in other to avoid birth complications.

According to a study done by Bartholomew C.O, Cyril C.I, et al. on Birth Preparedness and Complication Readiness among Pregnant Women in a Secondary Health Facility in Abakaliki, Ebonyi State in the Southeastern part of Nigeria, only 44.9% of the study population was aware of Birth Preparedness, and only 36.9% was aware of the complications.

According to a descriptive cross-sectional study done by Kuteyi, Kuku, et al (2011) on “Birth Preparedness and Complication Readiness of pregnant women attending the three levels of health facilities in Ife Central Local Government Nigeria” in the Southwestern part of the country, amongst 401 pregnant women. 39.3% of respondents were not aware of any danger signs in the pregnancy, childbirth, and postpartum periods, and as much as 79.4% of respondents had no arrangements for a blood donor.

In response to this pertinent issue, our study proposes to investigate several factors that cause poor birth preparedness and look out for possible remedies for them; we plan to use inclusive participatory investigations on pregnant mothers that attend the antenatal care clinic in ESUT Teaching Hospital, Parklane as a way of creating appropriate awareness and therefore hope to decrease the incidence of maternal and neonatal mortality.

## General objectives

### Objectives

The study of assessments of birth preparedness and complication readiness [bpcr] among pregnant mothers attending antenatal clinic in south Parklane.

### Specific objectives

┐ To access the knowledge of birth preparedness and complications readiness among pregnant mothers attending antenatal clinics in such Parklane

┐ To assess the level of birth preparedness and complication among pregnant mothers attending the antenatal clinic in Esuth Parklane

┐ To determine the perception of birth preparedness and complication readiness among pregnant mothers attending antenatal in such Parklane

┐ To determine factors affecting the perception and practice of birth preparedness and complication readiness among pregnant mothers attending antenatal clinic in ESUTH Parklane.

### Research Questions

What is the level of knowledge of pregnant women attending the antenatal clinic on the back? What are the factors that hinder pregnant women’s practice of birth preparedness and complication readiness?

Does bpcr have any effect on the outcome of pregnancy in such Parklane?

What is the relevance of back to the outcome of the pregnancy and maternal mortality in the long run?

### Justification

High maternal mortality is a major problem in the developing world. Globally, about 287,000 mothers die each year because of problems related to pregnancy and childbirth. About 99% of this occurs in developing countries; sub-Saharan Africa [56%] and South Asia [29%] accounts for 85% of the global burden of maternal death. Nigeria is unarguably the most populous country in sub-Saharan Africa and indeed in Africa as a whole and the maternity rate is more than 15x higher in slow progress in low and middle-income countries is explained by the low coverage of maternal health care and the 3 delays to health-seeking behaviors of mothers4.

This study has relevance to the general public, nurse practitioners, and administrators. It will be significant to the general public as findings from this study will equip the women and the general public with invaluable information on birth preparedness and complication readiness, which will go a long way if applied to ensure better and safer maternal health for mothers and their infants.

This study will also be relevant to the nurse practitioners as they will be equipped with the findings from this study, which will enable them to adequately prepare the couples for birth and possibly avoid any complications that may arise.

Findings from this study can be utilized by nurse administrators who will ensure that the necessary environments are provided that will enable the nurse to oversee the adoption of birth preparedness and complication readiness.

## Chapter Two

### Literature Review

High maternal mortality is a major problem in the developing world and Nigeria is no exception. Although birth preparedness and complication readiness have been widely studied in the developing world (Africa, India), thus the increasing knowledge and awareness of BP/CR in parts of these countries, results have shown that there has been poor practice among pregnant mothers, especially in Africa6.

Women and newborns need timely access to skilled care during pregnancy, childbirth, and the post-partum/newborn period. However, too often, their access to care is impeded by delays — delays in deciding to seek care, delays in reaching care, and delays in receiving care5.

These delays have many causes, including logistic and financial concerns, unsupportive policies, and gaps in services, as well as inadequate community and family awareness and knowledge about maternal and newborn health issues.

In a cross-sectional descriptive study to find out birth preparedness and complication readiness among the community as well as health facilities of Rewa district of Madhya Pradesh, India by the department of community medicine (2008 —09), SS Medical College Rewa, m.p., India, it was revealed that gap exists between knowledge and skills of health care providers. Also, the study showed that the BP index in the study population was found to be lower (47.5%). BP/CR index was significantly high in above-poverty-line families, with higher educational levels. It also revealed that knowledge of danger signs (18.6%), knowledge of transportation services (18.6%), 1st-trimester registration (24.1%), and population saved money (44.2%) were low7.

In the study carried out on “Birth Preparedness Among Antenatal Clients”, Utisoet. Al. (2008) evaluated birth preparedness and complication readiness among antenatal care clients. The study was a descriptive cross-sectional study, at the antenatal care clinic at Kenyatta National Hospital, Nairobi, Kenya. Clients who their study revealed that over 60% of the respondents were counseled by health workers on various elements of birth preparedness. 87.3% of the respondents were aware of their expected date of delivery, 84.3% had set aside funds for transport to the hospital during labor and 62.9% had funds for emergencies. 67% of the respondents knew at least one danger sign in pregnancy while only 6.9% knew of three or more danger signs. 109% of the respondents did not have a clear plan of what to do in case of an obstetric emergency. Level of education positively influenced birth preparedness.

Attitude and level of preparedness among 300 consenting pregnant women in the Indore city, India 2010, 47.8% of mothers were found to be well-prepared, and the remaining 52.2% were less prepared. 10.

Kuteyi, Kuku, Lateef, Ogundipe, Mogbeyteren, and Banjo (2011)11 carried out a descriptive cross-sectional study on “birth preparedness and complication readiness of pregnant women attending the three levels of health facilities in Ife central local government, Nigeria” among 401 pregnant women. 39.3% of respondents knew no danger signs in pregnancy, childbirth, and the postpartum period; only 6.0% had adequate knowledge of obstetric danger signs without prompting; 84.8% and 78.3% of women respectively had identified a birthplace and began saving money for delivery; 79.4% of made no. arrangement for a blood donor.

A Cross-sectional, Multicentre study was carried out in Benin Central Hospital and the University of Benin Teaching Hospital (UBTH) Edo State among 300 women on “the concept of birth preparedness in the Niger Delta of Nigeria” (Ibrahim, Owo e ye and Wagbats o ma, 2013)12. Their study showed that 38% of the respondents revealed some level of awareness of birth preparedness however, there are statistically significant differences in the source of information, level of education, and the expression of danger signs (all p-value <0.005) among these groups of women. Most (40.4%) embraced birth preparedness because it allows for ease of delivery, child spacing (28.1%) and to avoid complications (23.7%); and the majority of respondents in UBTH plan to achieve these goals by savings (92.1%), which is statistically different from those respondents from cub (z 3.59; p = 0.000).

Agbodohu (2013)13 assessed the knowledge and practices of birth preparedness and complication readiness among expectant mothers in Accra specifically to determine the association between socio-demographic factors and birth preparedness and complication readiness.

A cross-sectional design was employed with a sample size of 400 expectant mothers in their third trimester at the Ridge Regional Hospital. Though many of the mothers (77.3%) were aware of the fact that they may need blood during labor only 16.4% of mothers had blood in the blood bank and 31.6% said they had arranged for a blood donor. It was observed that almost two-thirds of the respondents knew some danger signs and gave one or two examples. Two-thirds did not know anything about eclampsia or pregnancy-induced hypertension. Almost all respondents (96%) had identified a close family member as a companion when in labor.

A study to assess the knowledge and practices towards birth preparedness and complication readiness and associated factors among women of reproductive age group in Ethiopia reveals poor knowledge and practices of birth preparedness and complication readiness. (Muhammedawel & Mesfin, 2014)15.

Kaso and Addisse (2014)16 also assessed knowledge and practices towards birth preparedness and complication readiness and associated factors among women of reproductive age group (15–49) in Robe Woreda, Arsi zone, Oromia region, Ethiopia.

Community-based cross-sectional study supplemented by qualitative design was conducted in January 2012. A total of 575 women were selected taking into account place of delivery identification, means of transportation, skilled attendant identification, and saving money, about 16.5% of the respondents were prepared for birth and its complications.

The study by August, Pembe, Kayombo, Mbekenga, Axemo, and Darj (2015) explored the perceptions, experiences, and challenges the community faced on bp/cr. A qualitative study design using focused group discussions was conducted. Qualitative content analysis was used to analyze the data.

The community members expressed a perceived need to prepare for childbirth. They were aware of the importance of attending the antenatal clinics, and relied on family support for practical and financial preparations such as saving money for costs related to delivery, moving closer to the nearest hospital, and also to use traditional herbs, in favor of a positive outcome. The community recognized that pregnancy and childbirth complications are preferably treated at the hospital. Facility delivery was preferred; however, certain factors including stigma on unmarried women and transportation were identified as hindering birth preparedness and hence utilization of skilled care. Challenges were related to the consequences of poverty, though maternal health care should be free, they perceived difficulties due to informal user fees.

Tobin, Ofili, Enebeli, and Enueze (2017)20 assessed BP/CR among pregnant women attending antenatal care in primary health care centers in Oredo Local Government area (LGA) in Benin City, Edo State. 49.6% were aware of at least one danger sign associated with pregnancy, labor, and postpartum, while 87.4% had identified a skilled birth attendant. 11.3% had saved money for obstetric care, and 62.2% had purchased or made plans to purchase birth supplies. 87.4% of respondents were found to be well-prepared for the birth. Having a tertiary education and being married were factors found to be significantly associated with BP. The majority of the women had BP/CR in place.

A study was done by Chioma Rose Nkwocha, Omosivie Maduka, and Faith C Diorgu from the Department of Nursing Science, University of Port Harcourt, Nigeria April (2017)21, among 407 consenting women. More than half of the pregnant women in the study 79 .12 % know about birth preparedness and complication readiness. The majority of the women 81.8% obtained the information from their healthcare providers. The practice of birth preparedness and complication readiness in the respondents showed that 93.61%had identified a place of delivery, 84.03% were saving money in case of emergencies, 89.93% were preparing essential items for safe delivery and the post-partum period, and 58.23% could detect early signs of an emergency. Birth preparedness and complication readiness knowledge and practice among the respondents were found to be satisfactory. The study noted that pregnant women rely so much on information provided by their healthcare providers during antenatal periods.

Kakaire, Kaye, and Osinde (2011) carried out a cross-sectional study on “male involvement in birth preparedness and complication readiness for emergency obstetric referrals in rural Uganda” among 140 women admitted as emergency obstetric referrals in antenatal, labor, or the postpartum period at the maternity ward of Kabale Regional Hospital, Uganda. Data was collected on socio-demographics and birth preparedness and what roles spouses were involved in during the development of the birth plan. Any woman who attended antenatal care at least 4 times, received health education on pregnancy and childbirth danger signs, saved money for emergencies, made a plan of where to deliver from, and made preparations for a birth companion, was deemed as having made a birth plan.

Multivariate logistic regression analysis was conducted to analyze factors that were independently associated with having a birth plan. The results of their study showed that concerning the role played by husbands in birth preparedness, sixty-two women (44.3%) made savings for an eventuality such as pregnancy complications, 60 (42.9%) were accompanied by the husband to the antenatal clinic, while 65 (43.4%) were accompanied to the labor ward by the husbands. It also revealed that on the factors associated with having a birth plan, the mean age was 26.8 ± 6.6 years, while the mean age of the spouse was 32.8 ± 8.3 years. Over 100 (73.8%) women and 75(55.2%) of their spouses had no formal education or only a primary level of education respectively.

On multivariable analysis, primigravidae compared to multigravidae (odd-ration (or) 1.8; 95°0ci 1.0-3.0), education level of the spouse of secondary or higher versus primary level or none (or = 3.8; 95%ci = 1.2 - 11 .0), formal occupation versus informal occupation of spouse (or = 1.6; 95%cl = 1.1 - 2.5), presence of pregnancy complications (or = 1.4; 95%ci = 1.1 -2.0) and the anticipated mode of delivery of cesarean section versus vaginal delivery (or = 1.6; 95%cl = 1.0 - 2.4) were associated with having a birth plan.

## Chapter Three

## Research Method

### 3.1. Study area

Enugu state is one of the 36 states of Nigeria, located in the southeastern part created in 1991 from the old Anambra State. It has a population of about 722,664 according to the 2006 Nigerian census.

Its capital and largest city is Enugu from which the state drives its name. The state is located at longitude 7.548949 and latitude 6.459964. The average temperature in the city is cool to mild; (60of) in its cooler months and gets warmer to hot in its warmer to hot in its warmer months (80of) and very good for outdoor activities.

The lowest rainfall of about 0.16cm3 (0.0098cu in) is normal in February while the highest is about 35.7cm3 (2.18cu in) in July.

Economically, the state is predominantly rural and agrarian, with a substantial portion of its working population engaged in farming. A small portion of the population is also engaged in manufacturing activities with the most pronounced among them located in Enugu, Oji, Ohebedim, and Nsukka.

Enugu State University Teaching Hospital College of Medicine is located at Park Avenue GRA Enugu North Local Government Area; the college of medicine is located inside the hospital. The Department of Obstetrics and Gynecology is located inside the hospital, having two (2) units: obstetrics and gynecology; they also have antenatal care, family planning, postnatal, and other sections.

### 3.2. Study design

This is a questionnaire-based cross-sectional descriptive study on the knowledge, attitude, and practice of birth preparedness and complication readiness among pregnant women who attend the antenatal care clinic in ESUT Teaching Hospital Parklane and will be based on volunteering anonymity and self-reporting.

### 3.3. Study population

The study will be conducted in ESUT teaching hospital Parklane using those who attend the antenatal care clinic.

#### 3.3.2. Exclusion criteria

1. Those not present on the day of examination
2. Those unwilling to participate.

### 3.4. Sampling determination

A sample size of 264 will be determined, the sample size was worked out using the Cochran formulae for determining the sample size.

**N=z2p q/d2.**

Where n, is the minimum sample size.

Z is the standard normal deviation usually set at 1.96 (or more at 2.0), which corresponds to the 95% confidence level.

P is 22% from prevalence studies. Q = 1.0 – p (22%)

But; p = 22/100 = 0.22 Therefore; q= 1 – 0.22 = 0.78

D is the degree of accuracy desired, usually set at 0.05 (constant)

**N = z2pq d2**

**N = 1.922 x 0.22 x 0.78 0.052**

**N = 1.922 x 0.22 x 0.78 25 x 10-4**

**= 263.688**

**N = 264**

### 3.5. Sampling technique

The sampling technique that will be used for the study is simple random sampling. All the pregnant women will have equal chances of being selected for the study, provided they meet the inclusion criteria.

### 3.6. Study instrument

The instrument that was used for data collection is a structured self-administered questionnaire. The study questionnaires were first presented and suitable modifications were done if any. The information that was collected includes demographic data of the respondents, awareness or knowledge, and practice of birth preparedness and complication readiness.

Data was collected in the antenatal care clinic. Appropriate instruction before filling out the questionnaire was given.

### 3.7 Method of data collection

Data collection was through the use of structured questionnaires.

### 3.8. Data analysis is

All analyses were performed using the statistical package for social science (spss) version 25.

The results were presented in tables and charts.

### 3.9. Ethical consideration

Ethical approval will be obtained from the research ethical committee of such teaching hospital Parklane.

administrative permission to carry out this study will be obtained from the hospital and presented to the head of the Department of Obstetrics and Gynecology. Written informed consent will be obtained from the respondents after the purpose of the study has been clearly explained to their understanding. Participation will be voluntary. Respondents will be free to withdraw from the study at any time and this will be properly communicated to them.

### 3.10. Anticipated limitations

- The pregnant women coming for antenatal care clinics in such teaching hospital parkland may not be fully cooperative.

**Table 1:** socio demographic.

| Variable | Frequency | Percentage |
| --- | --- | --- |
| 15- 20yrs | 12 | 4.5 |
| 21 - 30yrs | 159 | 60.2 |
| 31 - 40yrs | 92 | 34.8 |
| >40yrs | 1 | .4 |
| Total | 264 | 100.0 |
| **Marital status** |  |  |
| Single | 7 | 2.7 |
| Married | 257 | 97.3 |
| Total | 264 | 100.0 |
| **Highest educational level** |  |  |
| No formal education | 4 | 1.5 |
| Primary | 3 | 1.1 |
| Secondary | 43 | 16.3 |
| Tertiary | 214 | 81.1 |
| Total | 264 | 100.0 |
| **Religion** |  |  |
| Christian | 263 | 99.6 |
| Moslem | 1 | .4 |
| Total | 264 | 100.0 |
| **Ethnicity** |  |  |
| Igbo | 256 | 97.0 |
| Hausa | 3 | 1.1 |
| Yoruba | 5 | 1.9 |
| Total | 264 | 100.0 |

Most of our respondent are within the age range of 21-30 yrs 159 (60.2%), and majority are married 257 (97.3%), and the level of education with the highest respondent is tertiary 214 (81.1%) with many christians 263 (99.6%), and of igbo tribe 256 (97.0%).

**Table 2:**
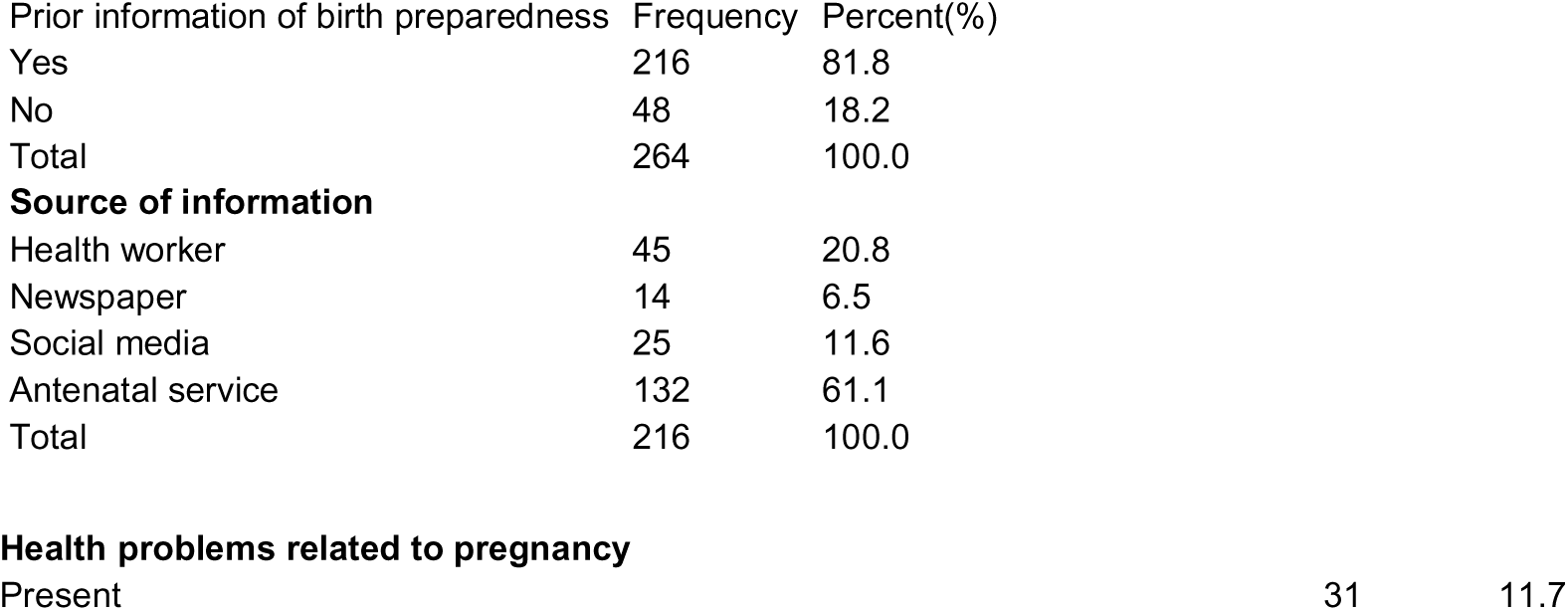
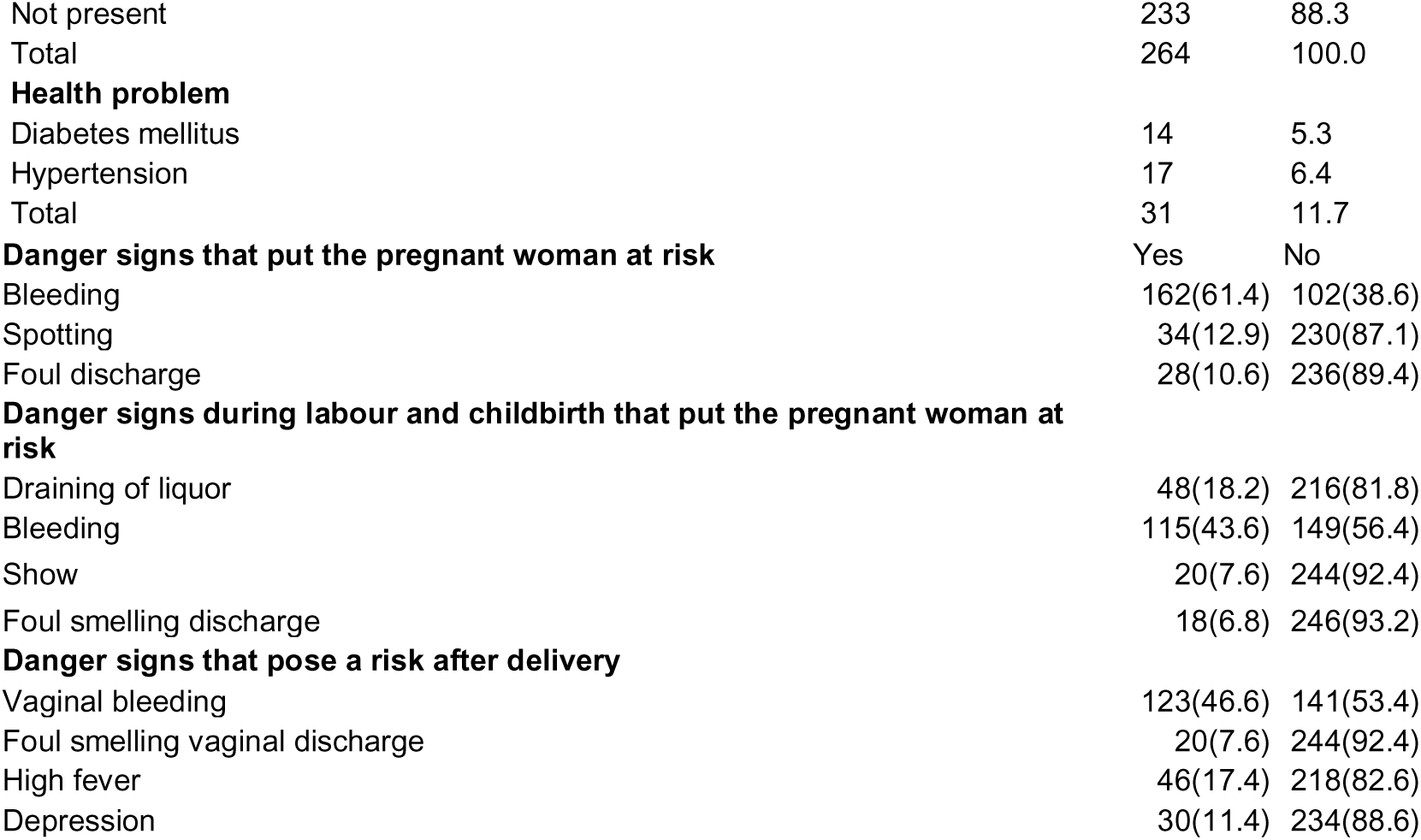
knowledge of birth preparedness and complication readiness.

About 216 (81.8%) of respondent had a good knowledge about birth preparedness and complication readiness and 132 (61.1%) got their knowledge during antenatal visits. About 162 (61.4%) had a fair knowledge of danger signs in pregnancies, 115 (43.6%) had a poor knowledge of danger signs of labor, and 123 (46.6%) had a poor knowledge on dangers after delivery.

**Table 3:** perception of birth preparedness and complication readiness.

| **Importance of birth preparedness and complication readiness** | Frequency | Percentage |
| --- | --- | --- |
| Important | 246 | 93.2 |
| Not important | 18 | 6.8 |
| Total | 264 | 100.0 |
| **Effect of regular antenatal visit on the outcome of your pregnant** |  |  |
| Effective | 205 | 77.7 |
| Not effective | 59 | 22.3 |
| Total | 264 | 100.0 |
| **Preference of work/home to your antenatal visit** |  |  |
| Work/home preferred | 29 | 11.0 |
| Work/home not preferred | 235 | 89.0 |
| Total | 264 | 100.0 |
| **Compliance to drugs regularly as prescribed** |  |  |
| Compliant | 208 | 78.8 |
| Not compliant | 56 | 21.2 |
| Total | 264 | 100.0 |
| **necessity of arranging a place for delivery** |  |  |
| Necessary | 251 | 95.1 |
| Not necessary | 13 | 4.9 |
| Total | 264 | 100.0 |
| **Place of preference for delivery** |  |  |
| Home | 14 | 5.6 |
| Mothers house | 1 | .4 |
| Midwives | 12 | 4.8 |
| Traditional birth attendant | 2 | .8 |
| Health centre | 11 | 4.4 |
| Teaching hospital | 211 | 84.1 |
| Total | 251 | 100.0 |
| **Need for government to provide adequate referral system** |  |  |
| Needful | 237 | 89.8 |
| Not needful | 27 | 10.2 |
| Total | 264 | 100.0 |

246 (93.2%) feels bpcr is important, 205 (77.7%) feels regular antenatal visits will have a positive effect on pregnancy outcomes. 29 (11%) prefers to stay at home/work than antenatal visits, 208 (78.8%) are compliant with their medications, and 251 (91%) 0f respondent feels it’s necessary to make arrangement for delivery. But however few prefers to deliver at home 14 (5.6%), midwives 12 (4.8%), 211 (84.1%) in a teaching hospital. Majority 237 (89.8%) feels the government should provide adequate referral system.

**Table 3:** preparation level of the index pregnancy.

|  |  |  |
| --- | --- | --- |
| **Place of preference for delivery** |  |  |
| Home | 5 | 2.0 |
| Mothers house | 7 | 2.8 |
| Midwives | 8 | 3.3 |
| Traditional birth attendant | 1 | .4 |
| Health centre | 13 | 5.3 |
| Teaching hospital | 212 | 86.2 |
| Total | 246 | 100.0 |
| **preparedness for this pregnancy** |  |  |
| Prepared | 241 | 91.3 |
| Not prepared | 23 | 8.7 |
| Total | 264 | 100.0 |
| **Emotions towards the pregnancy** |  |  |
| Happy | 219 | 83.0 |
| Sad | 1 | .4 |
| Not sure | 44 | 16.7 |
| Total | 264 | 100.0 |
| **routine drugs** |  |  |
| Yes | 245 | 92.8 |
| No | 19 | 7.2 |
| Total | 264 | 100.0 |
| **Do you keep to your antenatal appointments** |  |  |
| Yes | 250 | 94.7 |
| No | 14 | 5.3 |
| Total | 264 | 100.0 |
| **spouse's support** |  |  |
| Yes | 252 | 95.5 |
| No | 12 | 4.5 |
| Total | 264 | 100.0 |
| **savings for the arrival of the baby** |  |  |
| Yes | 218 | 82.6 |
| No | 46 | 17.4 |
| Total | 264 | 100.0 |
| **means of transportation to place of delivery** |  |  |
| Yes | 207 | 78.4 |
| No | 57 | 21.6 |
| Total | 264 | 100.0 |
| **Possible blood donor** | Frequency |  |
| Yes | 46 | 17.4 |
| No | 218 | 82.6 |
| Total | 264 | 100.0 |

Most of our respondent 241 (91.3%) are ready for their index pregnancy, 46 (17.4%) have identified a blood donor, 218 (82.6%) have saved money for the arrival of the baby, and about 207 (78.4%) have identified means of transport with 252 (95.5%) having a supportive spouse and moreover 219 (83%) are happy to be pregnant.

**Table 3b:** factors affecting birth preparedness and complication readiness.

|  |  |  |
| --- | --- | --- |
| **Rating of the antenatal services rendered to you** |  |  |
| Very bad | 4 | 1.6 |
| Bad | 11 | 4.5 |
| Good | 112 | 45.9 |
| Very good | 117 | 48.0 |
| Total | 244 | 100.0 |
| **Current family size** |  |  |
| 0 – 4 | 160 | 72.7 |
| 5 – 10 | 52 | 23.6 |
| Greater than 10 | 8 | 3.6 |
| Total | 220 | 100.0 |
| **monthly income** |  |  |
| Less than 5000 | 44 | 22.4 |
| 5000 – 50000 | 91 | 46.4 |
| 60000 – 100000 | 29 | 14.8 |
| Greater than 100,000 | 32 | 16.3 |
| Total | 196 | 100.0 |
| **husband's occupation** |  |  |
| Unemployed | 7 | 2.9 |
| Business man | 95 | 39.9 |
| Workman | 25 | 10.5 |
| Servant | 62 | 26.1 |
| Public servant | 20 | 8.4 |
| Professional | 29 | 12.2 |
| Total | 238 | 100.0 |
| **husband's monthly income** |  |  |
| Less than 5000 | 20 | 10.8 |
| 5000 – 50000 | 49 | 26.5 |
| 60000 – 100000 | 55 | 29.7 |
| Greater than 100,000 | 61 | 33.0 |
| Total | 185 | 100.0 |
| **Outcome of previous pregnancies** |  |  |
| Very satisfactory | 84 | 37.3 |
| Satisfactory | 40 | 17.8 |
| Less satisfactory | 22 | 9.8 |
| I don't know | 79 | 35.1 |
| Total | 225 | 100.0 |
| **Previous miscarriage** |  |  |
| Yes | 70 | 26.5 |
| No | 194 | 73.5 |
| Total | 264 | 100.0 |

About 117 (48%) of respondent are happy with the antenatal services rendered to them with 0-4 being the highest number of range of family size 160 (72.7%), and majority 91(46.4%) having a monthly income range of 5,000-50, 000. Most spouse are businessmen 95 (39.9%) with the highest monthly income range 61 (33.0%) > 100,000. About 84 (37.3%) had a very satisfactory prevous pregnancy outcome with majority of no previous miscarriage 194 (73.5%).

| Knowledge | Perception | Frequency (%) | Chi square |
| --- | --- | --- | --- |
| Prior knowledge of birth preparedness | Necessity to keep antenatal visit | 78.4 | 0.004 |
| Prior knowledge of birth preparedness | adequate referral by the government | 89.7 | 0.56 |
| Knowledge | Practice | Frequency (%) | Chi square |
| Prior knowledge of birth preparedness and complication readiness | Keeping to antenatal service | 94.7 | 0.004 |
| Prior knowledge of birth preparedness and complication readiness | Routine drugs | 92.8 | 0.002 |
| Prior knowledge of birth preparedness and complication readiness | Saved money for delivery | 82.6 | 0.03 |

Prior knowledge gives a positive perception of necessity of keeping to antenatal visits and adequate referral system by the government to. Prior knowledge has a good effect on the practice such as keeping to antenatal visits and compliance with routine drugs with a significance of 0.004 and 0.002 respectively. No significant findings on effects of prior knowledge to saving money for the pregnancy.

| Factors | Frequency (%) | Chi square (prior knowledge) | Chi square (necessary to know about bp/cr) | Chi square (keeps antenatal appointments) |
| --- | --- | --- | --- | --- |
| Family size (0-4) | 56.3 | 0.3 | 0.01 | 0.7 |
| Spouse support | 78.4 | 0.5 | 0.002 | 0.001 |
| Halth issues with index pregnancy | 11 | 0.3 | 0.3 | 0.7 |
| Outcome of previous pregnancies |  |  |  |  |
| Monthly income (5000-50,000) | 48 | 0.008 | 0.2 | 0.4 |
| Level of education | 32.1 | 0.2 | 0.3 | 0.3 |
|  | 76.5 | 0.08 | 0.2 | 0.004 |

76.5% of our respondent with tertiary level of education and 78.4% of supportive spouse keeps to antenatal visits with a significant finding of 0.004 and 0.001. However there is no significant findings of other factors effect on the knowledge, perception and practice of birth preparedness and complication readiness. Nevertheless 78.4% of supportive spouse of our respondent believes its necessary to have more knowledge of birth preparedness and complication readiness with a signicant finding of 0.002

## Chapter 5

## Discussion

### 5.2. Socio-demographic characteristics

Our research had respondents in the age category of 21-30 years (60.20%) the maximum number of married women is 97.35%. Christians are 99.6% and Igbo ethnicity is 97%. The study identified that 98.5% had formal education. This is in line with the study by Multiset. al. (2008), in Kenya which showed level of education positively influenced birth preparedness. Also, in keeping with the study by Kaso and Addisse (2014) in Ethiopia that showed preparation for birth and its complications was higher among educated women.

### 5.2. Knowledge of birth preparedness and complication readiness

Knowledge is necessary in birth preparedness and complication readiness. It is an important factor that plays a role in the healthcare-seeking behaviors of individuals. Concerning our study to our study in which 216 (81.8%) of respondents have heard about birth preparedness and complication readiness. This contrasts with a study done at Benin Central Hospital and the University of Benin Teaching Hospital which showed that 38% of respondents had heard about it. The difference in this level of awareness is because the majority of our study population 250 (94.7%) keeps to their routine antenatal appointments with about 132 (61.1%) hearing it first at antenatal clinics. Knowledge is interconnected with health care-seeking behaviors such as regular tabs on appointments and saving for the arrival of the baby and it is in keeping with our study in which knowledge about bp/cr was high

Secondly, our study showed that the highest number of respondents first heard about birth preparedness and complication readiness from antenatal services was 132 (61.1%), followed by health workers 45 (20.8), then social media 25 (11.6%). This demonstrated that antenatal services and health workers need to make more effort to enlighten pregnant mothers on bp/cr and also social media has an important role to play in the level of awareness. Unlike the study carried out in Port Harcourt Teaching Hospital were highest respondent 333 (81.8%) obtained the information from their health care providers.

Furthermore, our study showed that the respondent knew at least one danger sign of which vaginal bleeding was ranking the highest in pregnancy 162 (61.4%), childbirth 115 (43.6%) and postpartum 123 (46.6%) followed by foul-smelling vaginal discharge 28 (10.6%), 18 (6.8%), and 20 (7.6%) respectively and other symptoms. When compared to the work done in Ife Central Local Government, three levels of health facilities only 24 (6.0%) had adequate knowledge of obstetric danger signs. Although there was no adequate knowledge or rather half knowledge on obstetric danger signs in our study population the difference was quite high. This is so because of inaccuracy or half information from the informant.

### 5.3. Perception towards BP/CR

Our research showed that a vast majority of the population felt that birth preparedness and complication readiness were important (93.2%), with many pointing out that they would prefer to come to antenatal care(89%) as against staying at home or going to work, the compliance level of these women to regular antenatal care visits is (94.7%) and many more are on routine drugs(92.8%), it is, however, worrisome that those that believe regular antenatal care visits usually has a positive and effective outcome towards their deliveries is (77.7% ) and only (78.8%) are compliant with taking their routine drugs but this value was expected to increase, because according to the study of the skilled initiative (FCI, 2013)6 antenatal care attendance at least four times during pregnancy is a key component of birth preparedness and complication readiness, so this calls for better understanding and appreciation of antenatal care by pregnant women;

The study also revealed that (95.1%) felt that it is necessary to make arrangements for the delivery, but only (84.1%) preferred a teaching hospital, while 100% had already identified birthplace, with (86.2%) preferring the teaching hospital; a contrast study birth preparedness and complication readiness in Ife, Nigeria (2011)12 showed only (78.3%) as having made arrangements for a birthplace; this is encouraging as the teaching hospital is more equipped and efficient in handling obstetrics cases and any complications that may arise.

There is a high call for our government intervention in providing an adequate referral system to improve birth preparedness and complication readiness in our society.

Therefore, this high perception shows that women attending antenatal care in ESUTH feel that it will be beneficial to them and will ultimately improve their chances of having a good pregnancy outcome.

### 5.4. Level of preparation among the study populace

In our study, (91.3%) are prepared for the birth of the child, with the majority having happy emotions (83%) towards the pregnancy. In contrast with our study in Indore City India 2010 where 47.8% were prepared for their pregnancy. This discrepancy may result from their acceptance of the education of BP/CR and greater effort in securing a good outcome evidenced by compliance with antenatal care appointments and routine drugs 94.4% and 92.8% respectively.

Spouse support was (95.5%), this is heartwarming due to the key role played by the male partner, especially in decision-making, this is to studies done by **(Singh, Darroch, Vlassoff, & Nadeau, 2003)35, (Yue, O’Donnell &, 2010)36 and(Gross, Alba & Gras s; 2012, Abose, Woldie & Ololo, 2010)38** they highlighted the role of men in the outcome of birth, impact on maternal health and reduction in maternal **deaths**, decision-making about family planning and use of contraceptives; respectively.

This is reflected in the number of respondents that have saved money for birth (82.6%), which is much higher than descriptive cross-sectional studies on BPCR done in; Kano, Nigeria(2010)9 showed that just 19.5% have money saved and in India by departments of community medicine(2008-2009)7, revealed that only (44.2%) have saved money; although in a similar study on concept of birth preparedness in the delta of Nigeria in UBTH (2013)13, showed a higher financial preparation than ours at (92.1%) having saved money.

Respondents who have identified means of transportation to a place of delivery is (78.4%), this is a key area for the spouse to show support as indicated in a study by Waiswa (2008)34; it was noticed that a vast majority of respondents have not identified any blood donor in case of emergency (82.6%), with only (17.4%) making arrangement and this is a big cause of concern since pregnancy is associated with many hematological conditions e.g bleeding or hemorrhages, blood loss from operative conditions, severe anemia, this is in contrast to a study done by Agbodohu (2003)14 in Accra, Ghana on BPCR, showed that while 31.6% has identified a blood donor, only 16.4% had blood in a blood bank.

### 5.5. Factors affecting preparation level among pregnant mothers

Respondents that have the highest value in family size was between 0-4 (72.7%), showing that the majority of respondents are relatively young and inexperienced families; this might affect the level of preparedness “fear of the unknown for the young inexperienced mothers” and “I know it all for the experienced multiparous” causing relative effective consciousness and preparedness and ineffective preparedness respectively. The financial power of the husbands (33% earning above 100,000naira) is greater than the women (46.4% earning 5,000-50,000naira), and this will affect the decision-making of women causing less preparation. This represents another key role the spouse will play.

An interesting finding in this study is that only (37.3%) had a very satisfactory outcome in a previous pregnancy, with 26.5% having had a miscarriage, this is worrisome in that it exposes the increased risk of maternal and infant mortality in our society, therefore greater care must be advocated from the pregnant woman to the medical staffs, down to the government to ensure the higher satisfactory outcome in future deliveries.

Kakaire, Kaye, and Osinde (2011) 11, believed that any woman who attended antenatal care at least 4 times, received health education on pregnancy and childbirth danger signs, saved money for emergencies, identified a blood donor made a plan of where to deliver and made preparations for a birth companion, was deemed as having made a birth plan. This should positively affect the level of preparation and complication readiness.

## Chapter 6

## Conclusion

Knowledge of birth preparedness is very high among the study population, although knowledge of complication signs and readiness is poor due to differences in the knowledge of informants. A high level of spouse involvement in our study population and compliance to antenatal visits and routine drugs, good antenatal services, stable financial income, family size, and outcome of previous pregnancies affect the level of preparation and complication readiness and should lead to good pregnancy outcomes, improved family health, increase living status and reduced maternal and infant mortality rate.

## Recommendations

Based on our findings in our study, we suggest the following;

1. The government at all levels, should be incorporated in the system of bp/cr employing an adequate referral system
2. Education on birth preparedness and complication readiness should be incorporated and improved on in the antenatal visits schedule.
3. Our social media (television, telecommunications, newspaper, and social networks (Facebook, Instagram, Twitter, etc)), should be involved in the dissemination of the correct information.
4. Health workers and all enlightened should be equipped to disseminate correct and relevant information on BP/CR.
5. Birth preparedness and complication readiness should be incorporated into the curriculum of schools and universities thereby increasing the scope of participation
6. Birth preparedness and complication readiness should be incorporated into the agendas of the Ministry of Health, government hospitals, teaching hospitals, private hospitals as well as primary health care centers. This will encourage the practice of this.
7. Encourage male participation like husbands attending antenatal visits with their wives and also teach them birth preparedness and complication readiness because most time husbands are observant.

## Data Availability

All data produced in the current work are present in the manuscript.

## Questionnaire

## Introduction

we are 5th-year medical students at the Esut College of Medicine researching the knowledge, attitude, and practice of birth preparedness and complication readiness among women attending antenatal in the Enugu metropolis. To achieve this, your participation and co-operation are highly needed. Responses to this questionnaire indicate consent and will be treated with absolute confidentiality.

kindly tick only one response for each question or information required except where otherwise instructed. Where blank spaces exist, kindly fill them with appropriate answer(s) Thanks for your cooperation

**Section a: demographic data**

1. Age: 15-20 [ ] 21-30[ ] 31-40[ ] >40[ ] Specify
2. Martial status: single [ ] married [ ] divorced [ ] widowed [ ] separated [ ]
3. Highest educational level attained: no formal education [ secondary[ ] (d) tetiary[ ]
4. Religion: Christian [ ] (b) Muslim [ ] (c) traditionalist [ If others, specify
5. Ethnicity: Igbo [ ] (b) Hausa [ ] (c) yoruba [ ] If others, specify ] (b) primary [ ] ] (c)

**SECTION B: KNOWLEDGE OF BIRTH PREPAREDNESS AND COMPLICATION READINESS**

Have you ever been pregnant? Yes [ ] no [ ]

Have u heard of birth preparedness and complication readiness yes[ ] no[ ]

Source of information: health worker[ ] (b) newspaper[ ] (c) social media[ ] antenatal service[ ]. If others, specify……

Any personal health problem related to pregnancy. Yes [ ] no [ ]

If yes answer the next question

Health problem: diabetes mellitus [ ] (b) hypertension [ ](c) sickle cell. If others please specify……………..

Danger signs that put the pregnant woman at risk: bleeding[ ] (b) spotting[ ] (c) foul discharge[ ]

If others, please specify………

Danger signs during labor and childbirth that put the pregnant woman at risk. (a)draining of liquor [ ] (b) bleeding [ ] (c) show [ ] (d) foul smelling discharge[ ]

If others, please specify……

Danger signs that pose a risk after delivery. (a) vaginal bleeding (b) foul-smelling vaginal discharge (c) high fever (d) depression

**section c: perception towards birth preparedness and complication readiness**

Do you think it’s necessary to know about birth preparedness and complication readiness? Yes [ ] no [ ]

Do you feel regular antenatal visits will affect the outcome of your pregnancy? Yes [ ] no [ ]

Would you rather stay at work/home than go to your antenatal visit? Yes [ ] no [ ]

Do you take your drugs regularly as prescribed? Yes [ ] no [ ]

Do you feel it’s necessary to arrange for your delivery? Yes [ ] no[ ]

Where? Home [ ] (b) mother house [ ] (c) midwives [ ] (d) traditional birth attendant[ ] (e) health center [ ] (f) teaching hospital [ ]

Do you think it should be taught to other people around? Yes [ ] no [ ]

Do you think the government should provide an adequate referral system? Yes [ ] no[ ]

Section c: **factors affecting birth preparedness and complication readiness.**

Are you currently pregnant? Yes [ ] no [ ]

Where is your preferred delivery place? (a) home [ ] (b) mother house [ ] (c) midwives [ ] (d) traditional birth attendant[ ] (e) health center [ ] (f) teaching hospital [ ]

Are you prepared for this pregnancy? Yes [ ] no [ ]

how do you feel about the pregnancy? (a) happy (b)sad (c) no T sure

Are you on your routine drugs? Yes [ ] no[ ]

Do you keep to your antenatal appointments? Yes [ ] no [ ]

Is your spouse supportive? Yes [ ] no [ ]

Have you saved money for the arrival of the baby? Yes [ ] no[ ]

Have you identified means of transportation to the place of delivery? Yes [ ] no [ ] Have you identified any blood donors? Yes [ ] no [ ]

Rate the antenatal services rendered to u? (a) very bad (b)bad (c) good (d) very good

what’s your family size? (a) 0-4 [ ] (b) 5-10 [ ] (c) greater than 10 [ ]

What’s your monthly income? (a) less than 5000 (b) 5000-50,000 (c) 60,000-100,000 (d) greater than 100,000

What’s your husband’s occupation? (a) unemployed (b) businessman (c) workman (d) civil servant (e) public servant (f) professional

If others, please specify……..

What’s your husband’s monthly income? (a) less than 5000 (b) 5000-50,000 (c) 60,000-100,000 (d) greater than 100,000

The outcome of previous pregnancies: (a) very satisfactory (b) satisfactory (c) less satisfactory (d) I don’t know

Any previous miscarriage? Yes [ ] no [ ]

## Notes

### Competing Interest Statement

The authors have declared no competing interest.

### Funding Statement

This study did not receive any funding.

### Author Declarations

Ethical committee of Enugu State University Teaching Hospital Parklane gave ethical approval for this work.

